# Drivers of youth mental health and wellbeing: A nationwide cross-sectional study in Morocco

**DOI:** 10.1101/2025.03.28.25324837

**Authors:** Jihad Bnimoussa, Oumnia Bouaddi, Imad Elbadisy, Mohamed Khalis

## Abstract

**Background:** Mental health struggles disproportionately affect young people, particularly in low- and middle-income settings. Understanding key drivers of youth mental health is essential for designing effective policies and programs to close the mental health gap. This study aims to describe the factors influencing mental health and well-being among Moroccan youth.

**Methods:** This is a descriptive cross-sectional online survey conducted across 12 regions in Morocco, using stratified random sampling. Moroccan youth aged 18 to 24 were included. Data were collected using a questionnaire informed by and adapted from the WHO Adolescent Wellbeing Framework, distributed through youth-serving organizations.

**Results:** A total of 1,182 participants were included (mean age 20.5 years, 68.2% female, 85.7% from urban settings). Regarding health and nutrition, 46.3% valued sleep, 59.7% emphasized physical health, 53.1% highlighted access to quality healthcare, and 56.5% prioritized clean air. In terms of connectedness and contribution, 75.7% rated family relationships as critical to their well-being, while 42.5% emphasized positive peer relationships. Regarding safety and supportive environments, 64.7% considered personal safety essential, 70% prioritized the fulfillment of basic needs, and 63.7% valued personal information protection. For education and competence, 54.4% emphasized learning opportunities and 62.2% identified self-confidence as key drivers. Regarding agency and resilience, 59.4% valued independence, 68.5% stressed having a sense of purpose, and 55% identified hope and optimism as key to their well-being. In digital well-being, 37.7% believed social media helped maintain connections, 38% viewed it as a learning tool, while 31.6% reported it as a source of stress and anxiety.

**Conclusion:** This study provides valuable insights into priority drivers of youth mental health in Morocco which should be the target for future interventions aiming to promote youth well-being. The findings contribute to the limited data on youth mental health in LMICs, highlighting the urgency for comprehensive mental health services and further research.

## 1. Background

Mental health struggles are a critical global issue, disproportionately affecting young people who bear a significant burden of these challenges (1). Adolescents, particularly those aged 15 to 25, are highly susceptible to mental health problems due to neurological and psychosocial changes. Conditions such as depression, anxiety, eating disorders, and self-harm can severely impact their development, academic performance, social relationships, and future outlook. The consequences of these issues can be severe, including self-harm and suicide, which is the third leading cause of mortality among youth globally (1). Low to Middle Income countries face disproportionate gaps in their youth mental health systems and these landscapes of youth development create an additional challenge to addressing the unmet needs of young people (2).

Young people in low to middle-income countries (LMICs) account for 90% of the world’s youth population (3) who often experience inequities in access to resources and opportunities. The disparity in mental health care resources exacerbates the problem, with less than 2% of health care budgets being allocated to mental health (1). A critical issue is the shortage of mental health professionals in these regions, with an average of just one professional for every 200,000 people (1). While high-income countries generally have better access to psychiatric services, mental health professionals, and crisis response mechanisms, LMICs, like Morocco, suffer from a severe lack of infrastructure, with limited psychiatric facilities concentrated in urban areas and an unregulated private and non-governmental organizations (NGO) sector (4).

In Morocco, the private sector, comprising 233 psychiatrists and only 9 child psychiatrists (5), is largely concentrated in urban centers, making access to specialized care difficult for many (5). Non-governmental organizations (NGOs) and civil society organizations (CSOs) play a supplementary role, with a few associations offering support for specific conditions like autism and disabilities (6). Mental health issues among Moroccan youth are further compounded by widespread substance abuse, depression, anxiety, and other psychological disorders. The Mediterranean Survey on Alcohol and Other Drug Use in Schools (MedSPAD) (6) highlighted troubling rates of substance use, including tobacco (20% in boys, 6% in girls), cannabis (9.5% in boys), and psychotropic substances (4.4% in girls), starting as early as 15 years old (6). Moreover, the Global School-based Student Health Survey 2016 (7) revealed significant levels of depression, anxiety, and insomnia, with 16.8% of students reporting sleep disturbances due to worry. Suicidal thoughts and behaviors are alarmingly prevalent, with 16% of urban students having seriously considered suicide (7). The current literature on what drives youth mental health and wellbeing in Morocco is limited and consists of small-scale studies largely focusing on accessible groups such as students (8–11). Despite efforts to prioritize youth mental health and wellbeing and increased interest in the area, the current evidence base is marked by significant gaps regarding the drivers of youth mental health and well-being. Thus, the aim of this study is to identify important drivers of mental health and well-being among Moroccan youth aged 18 to 24 across the 12 regions of Morocco. This study makes a significant contribution to youth mental health research in LMICs, particularly in Morocco, by providing national-scale data from a youth perspective and has important implications for research, programming, and policy initiatives aimed at addressing the unique challenges faced by young people in these regions.

## 2. Methods

### 2.1 Conceptual Framework

Mental Health, the state of mental wellbeing, can be divided into subjective and objective wellbeing, with each approach emphasizing different indicators of youth wellbeing. Subjective wellbeing highlights individual fulfillment and the lived experience of young people while the objective definition of wellbeing tends to focus on quality-of-life indicators (12). The expanded definition of wellbeing developed as part of the Adolescent Wellbeing Framework combines the two constructs: “Adolescents have the support, confidence, and resources to thrive in contexts of secure and healthy relationships, realizing their full potential and rights.”

The data collection survey used in this study was developed based on the Adolescent Wellbeing Framework (AWF), a framework developed by the Partnership for Maternal, Newborn and Child Health, the World Health Organization, and the UN H6+ partner agencies (12). The framework includes five components that determine adolescent wellbeing (Figure 1): (1) good health and optimum nutrition; (2) connectedness, positive values, and contribution to society; (3) safety and a supportive environment; (4) learning, competence, education, skills, and employability; (5) agency and resilience. While the original framework does not include assessment of young people’s digital life space, a sixth component, (6) digital wellbeing and literacy, was added to the survey, emergent from desk research and consultations with Moroccan youth, to explore the digital environment, virtual connectivity, and internet use as a wellbeing driver.

**Figure 1.**
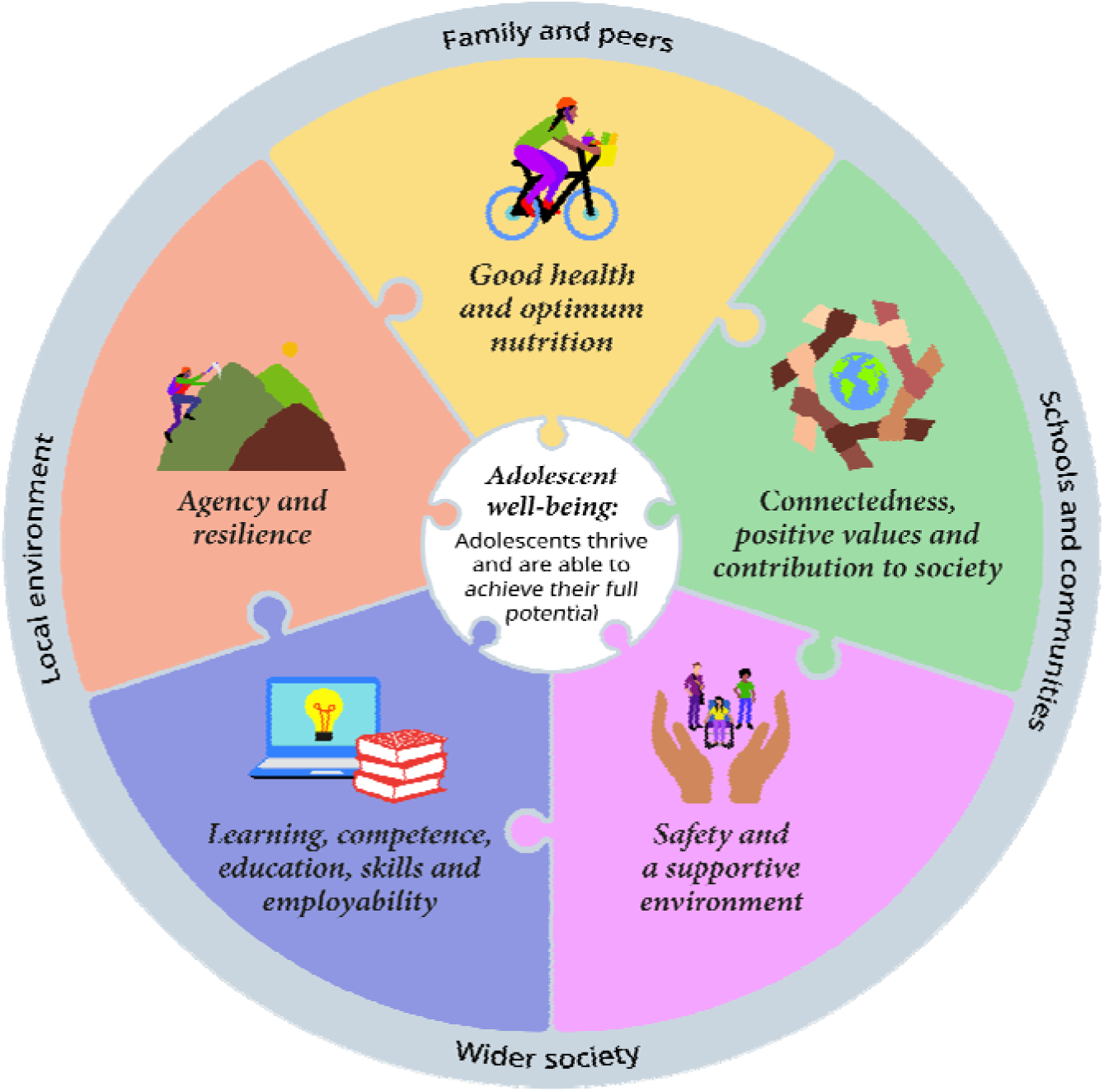
Adolescent Wellbeing Framework (12).

### 2.2 Study design and setting

A cross-sectional self-administered online survey was conducted among Moroccan young people between the ages of 18 and 24 years old across the 12 regions of Morocco. The study period took place between November 2023 and February 2024. The survey was distributed through local networks of youth serving organizations using WhatsApp, and was completed online, via Microsoft Forms, after informed consent.

### 2.3 Study population

In this study we included young people aged between 18 and 24 years old from varying socio demographic backgrounds residing in any of the 12 regions of Morocco. In order to include young people representing the 12 regions of Morocco, stratified random sampling was used to allocate sample size per regions and sub-regions, with sociodemographic characteristics such as place of residence, sex, education status being considered in participant outreach.

### 2.4 Data collection

Twelve young people were trained in participant recruitment and data collection, one for each region. These individuals were identified, selected, and trained for their work within civil society and community organizations that have an established network within the target population. Within each of the 12 regions, data was collected using a self-administered online survey, using MicrosoftForms, which was made available in both Arabic and French. The survey was developed by the study team and validated by experts, based on the Adolescent Wellbeing Framework (Supplementary material 1). For each item within the five domains, with each domain containing between 6 to 16 items, participants indicated the level of importance the item rates in their wellbeing using a 5-point Likert scale. The survey was piloted among 20 participants who met the inclusion criteria with feedback used to edit and modify the survey prior to its use for data collection. The English version of the questionnaire has been added to supplementary material.

### 2.5 Data analysis

Data collected from the online survey was collated into an excel spreadsheet and cleaned to ensure correct data entry. The total number of survey results used for analysis after data cleaning were 1182 results (n = 1182). Descriptive analysis was performed using the Jamovi 2.3.28 software. Quantitative variables were expressed in means and standard deviations, and qualitative variables were expressed in frequencies and percentages.

## 3. Results

### 3.1 Socio-demographic characteristics

A total of 1182 responses were collected and analyzed through local community networks across the 12 regions of Morocco (Table 1). The mean age of participants was 20.5 years old. A total of 68.2% of participants identified as female while only 31.8% were male. The majority of participants 85.7% were from an urban setting. 97.2% of participants reported their marital status as single. About 88.6% reported their highest education level being higher education or professional/vocational training with the majority of participants 81.7% being students at the time of the study.

**Table 1.**
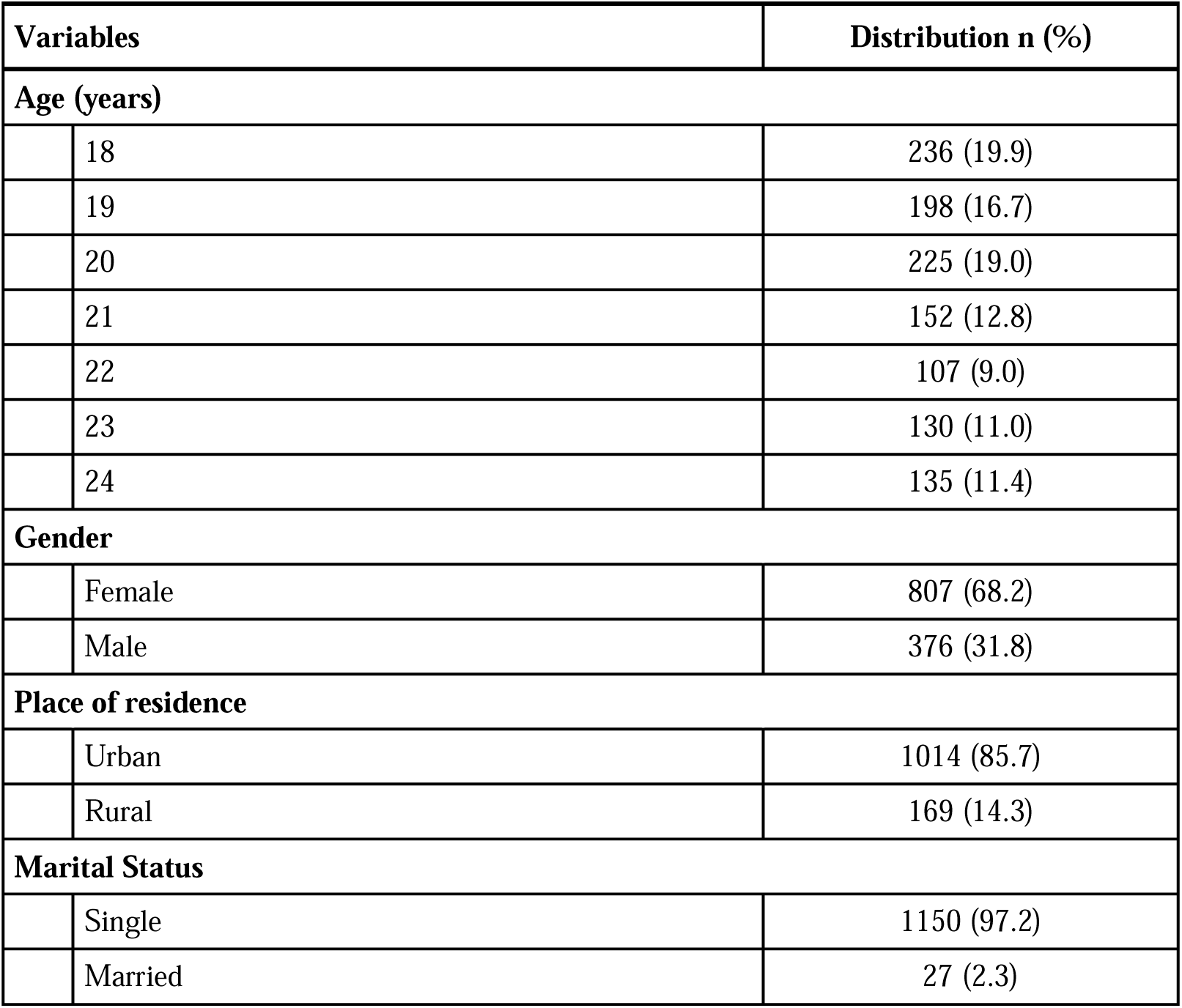
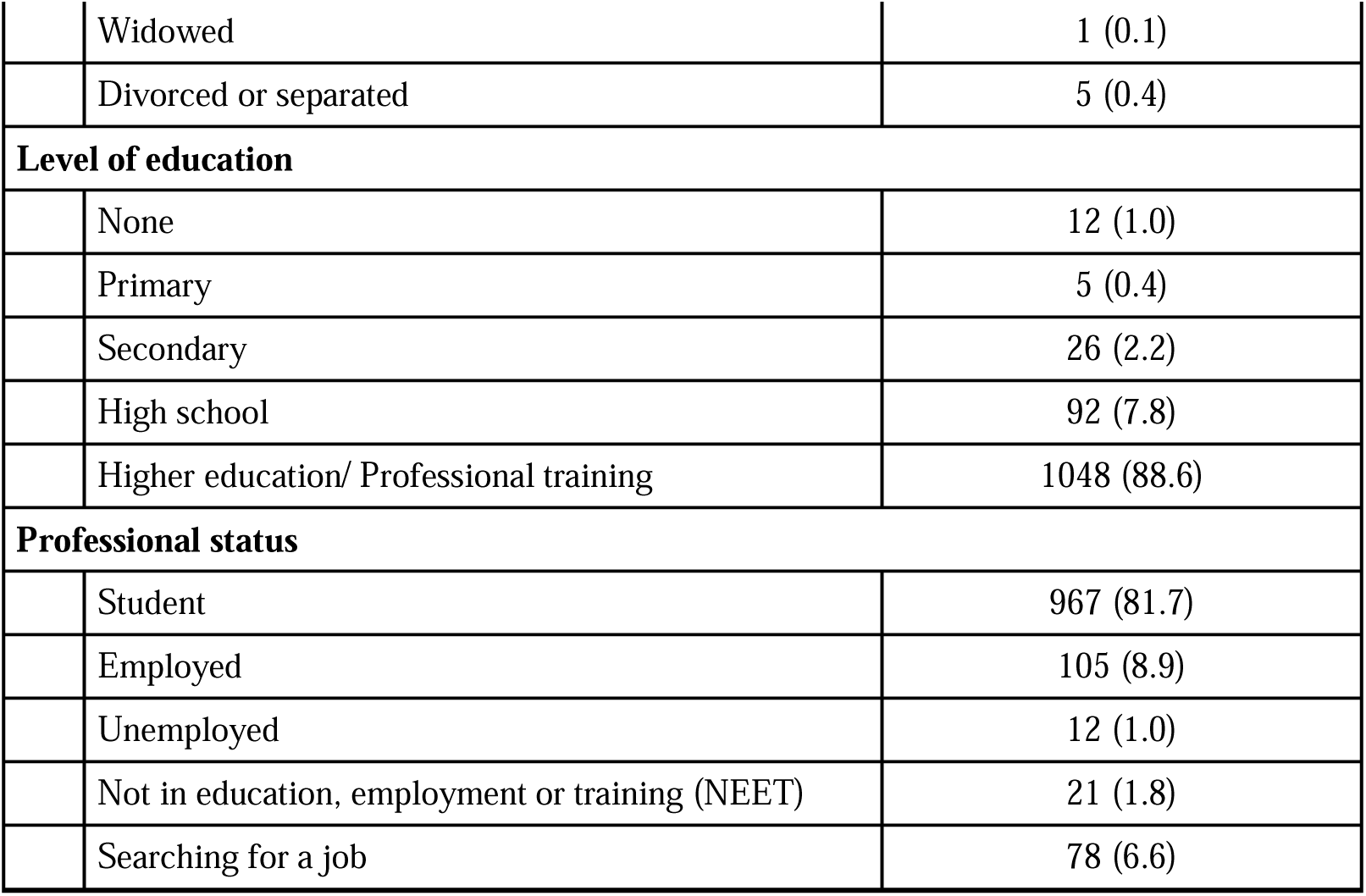
Sociodemographic characteristics of participants (*n* = 1182).

| Variables |  | Distribution n (%) |
| --- | --- | --- |
| **Age (years)** |  |  |
|  | 18 | 236 (19.9) |
|  | 19 | 198 (16.7) |
|  | 20 | 225 (19.0) |
|  | 21 | 152 (12.8) |
|  | 22 | 107 (9.0) |
|  | 23 | 130 (11.0) |
|  | 24 | 135 (11.4) |
| **Gender** |  |  |
|  | Female | 807 (68.2) |
|  | Male | 376 (31.8) |
| **Place of residence** |  |  |
|  | Urban | 1014 (85.7) |
|  | Rural | 169 (14.3) |
| **Marital Status** |  |  |
|  | Single | 1150 (97.2) |
|  | Married | 27 (2.3) |
|  | Widowed | 1 (0.1) |
|  | Divorced or separated | 5 (0.4) |
| **Level of education** |  |  |
|  | None | 12 (1.0) |
|  | Primary | 5 (0.4) |
|  | Secondary | 26 (2.2) |
|  | High school | 92 (7.8) |
|  | Higher education/ Professional training | 1048 (88.6) |
| **Professional status** |  |  |
|  | Student | 967 (81.7) |
|  | Employed | 105 (8.9) |
|  | Unemployed | 12 (1.0) |
|  | Not in education, employment or training (NEET) | 21 (1.8) |
|  | Searching for a job | 78 (6.6) |

### 3.2 Mental Health and Wellbeing Drivers

Table 2 presents how participants rated the importance of various drivers to their wellbeing (Table 2). Below we present the results for each component.

**Table 2.** Mental health and well-being drivers among young Moroccans (*n* = 1182).

| Variables | Distribution N (%) |  |  |  |  |
| --- | --- | --- | --- | --- | --- |
|  | Extremely important | Very important | Moderately important | Slightly important | Not important at all |
| **Good health and optimum nutrition** |  |  |  |  |  |
| Having a varied and balanced diet | 341 (28.8) | 469 (39.6) | 227 (19.2) | 85 (7.2) | 61 (5.2) |
| Sleeping well at night | 548 (46.3) | 425 (35.9) | 133 (11.2) | 37 (3.1) | 40 (3.4) |
| Practicing in sports and exercise | 319 (27.0) | 463 (39.1) | 234 (19.8) | 94 (7.9) | 73 (6.2) |
| Being satisfied with the way I look | 418 (35.3) | 468 (39.6) | 176 (14.9) | 64 (5.4) | 57 (4.8) |
| Being physically healthy | 706 (59.7) | 365 (30.9) | 61 (5.2) | 18 (1.5) | 33 (2.8) |
| Having access to information about health | 308 (26.0) | 494 (41.8) | 240 (20.3) | 83 (7.0) | 58 (4.9) |
| Having access to quality health services | 628 (53.1) | 374 (31.6) | 107 (9.0) | 30 (2.5) | 44 (3.7) |
| Having access to green spaces | 472 (39.9) | 442 (37.4) | 167 (14.1) | 60 (5.1) | 42 (3.6) |
| Breathing clean air | 664 (56.5) | 359 (30.5) | 94 (8.0) | 28 (2.4) | 31 (2.6) |
| **Connectedness, positive values, and contribution to society** |  |  |  |  |  |
| Having a good relationship with my parents and family | 896 (75.7) | 212 (17.9) | 39 (3.3) | 14 (1.2) | 22 (1.9) |
| Having a positive relationship with my teachers/ professors/ employer | 411 (34.7) | 483 (40.8) | 199 (16.8) | 49 (4.1) | 41 (3.5) |
| Having a positive relationship with my peers/ colleagues | 297 (25.1) | 503 (42.5) | 267 (22.6) | 73 (6.2) | 43 (3.6) |
| Feeling a sense of belonging in my community, school or work environment | 396 (33.5) | 512 (43.3) | 172 (14.5) | 54 (4.6) | 49 (4.1) |
| Having an adult person in my life who I can trust | 390 (33.0) | 383 (32.4) | 230 (19.4) | 95 (8.0) | 85 (7.2) |
| Feeling accepted, respected and valued by others | 435 (36.8) | 424 (35.8) | 202 (17.1) | 65 (5.5) | 57 (4.8) |
| Participating in social and cultural activities in the community or at school /university or work | 335 (28.3) | 390 (33.0) | 264 (22.3) | 118 (10.0) | 76 (6.4) |
| Having the chance to participate in decision-making and having my ideas valued and respected. | 472 (39.9) | 442 (37.4) | 171 (14.5) | 51 (4.3) | 47 (4.0) |
| Getting the chance to learn about empathy, make friends, and become more understanding | 354 (29.9) | 461 (39.0) | 227 (19.2) | 85 (7.2) | 56 (4.7) |
| **Safety and a supportive environment** |  |  |  |  |  |
| Not being exposed to violence (including bullying, online harassment, physical, sexual, verbal abuse and emotional violence) | 704 (59.5) | 301 (25.4) | 75 (6.3) | 47 (4.0) | 56 (4.7) |
| Feeling safe in my daily life, whether at home, in my neighborhood, online, or at school or work. | 765 (64.7) | 302 (25.5) | 59 (5.0) | 26 (2.2) | 31 (2.6) |
| Being treated like others and without discrimination | 597 (50.5) | 368 (31.1) | 118 (10.0) | 46 (3.9) | 54 (4.6) |
| Feeling safe to express myself and be who I am | 630 (53.3) | 388 (32.8) | 93 (7.9) | 29 (2.5) | 43 (3.6) |
| Having essential needs such as food, water, a place to live, warmth, clothing, and feeling safe and secure | 828 (70) | 252 (21.3) | 54 (4.6) | 20 (1.7) | 29 (2.5) |
| My personal information is protected and is not shared without my permission | 754 (63.7) | 295 (24.9) | 74 (6.3) | 25 (2.1) | 35 (3.0) |
| Having access to leisure activities and personal development opportunities | 447 (37.8) | 449 (38.0) | 174 (14.7) | 68 (5.7) | 45 (3.8) |
| **Learning, competence, education, skills, and employability** |  |  |  |  |  |
| Getting to go to school and having chances to keep learning, whether in a classroom or through other kinds of learning | 643 (54.4) | 385 (32.5) | 94 (7.9) | 26 (2.2) | 35 (3.0) |
| Getting help to stay motivated and keep learning, | 436 (36.9) | 468 (39.6) | 173 (14.6) | 62 (5.2) | 44 (3.7) |
| Having chances to build the tools and skills to succeed | 552 (46.7) | 473 (40.0) | 103 (8.7) | 17 (1.4) | 38 (3.2) |
| Having self-confidence and feeling that I can do things well | 736 (62.2) | 333 (28.1) | 58 (4.9) | 17 (1.4) | 39 (3.3) |
| Learning practical skills for work | 614 (51.9) | 410 (34.7) | 96 (8.1) | 30 (2.5) | 33 (2.8) |
| Working in jobs and businesses appropriate for my age | 481 (40.7) | 407 (34.4) | 180 (15.2) | 53 (4.5) | 62 (5.2) |
| Being satisfied with my learning and skills | 609 (51.5) | 431 (36.4) | 84 (7.1) | 28 (2.4) | 31 (2.6) |
| Believing in myself and my ability to reach my learning goals | 708 (59.8) | 356 (30.1) | 54 (4.6) | 26 (2.2) | 39 (3.3) |
| **Agency and resilience** |  |  |  |  |  |
| Feeling independent and capable of making my own decisions | 703 (59.4) | 370 (31.3) | 71 (6.0) | 11 (0.9) | 28 (2.4) |
| Feeling empowered to accomplish things and having confidence in myself (whether with friends, family, or in decision-making) | 660 (55.8) | 406 (34.3) | 74 (6.3) | 14 (1.2) | 29 (2.5) |
| Having hope and optimism towards the future | 651 (55.0) | 407 (34.4) | 68 (5.7) | 24 (2.0) | 33 (2.8) |
| Having a sense of purpose in my life | 810 (68.5) | 263 (22.2) | 62 (5.2) | 14 (1.2) | 34 (2.9) |
| Having opportunities to develop the ability to handle the challenges in life both now and in the future | 622 (52.6) | 434 (36.7) | 78 (6.6) | 18 (1.5) | 31 (2.6) |
| Having chances to reach my full potential now and later in life. | 672 (56.8) | 379 (32) | 83 (7.0) | 17 (1.4) | 32 (2.7) |

| Digital well-being and literacy | Completely agree | Agree | Undecided | Disagree | Strongly disagree |
| --- | --- | --- | --- | --- | --- |
| Social media helps me stay connected with my friends and family. | 446 (37.7) | 570 (48.2) | 93 (7.9) | 47 (4.0) | 27 (2.3) |
| I feel pressure to portray a perfect life on social media. | 147 (12.4) | 222 (18.8) | 213 (18.0) | 281 (23.8) | 320 (27.0) |
| Social media has a positive impact on my self-esteem. | 131 (11.1) | 306 (25.9) | 340 (28.7) | 228 (19.3) | 178 (15.0) |
| Some people make fun of me on social media platforms which affects my well-being | 109 (9.2) | 175 (14.8) | 206 (17.4) | 232 (19.6) | 461 (39.0) |
| I find it difficult to disconnect from social media, even when I want to. | 235 (19.9) | 305 (25.8) | 243 (20.5) | 217 (18.3) | 183 (15.5) |
| Social media provides a platform for me to express my creativity and interests. | 244 (20.6) | 465 (39.3) | 249 (21.0) | 131 (11.1) | 94 (7.9) |
| Comparing myself to others on social media makes me feel inadequate. | 203 (17.2) | 275 (23.2) | 214 (18.1) | 185 (15.6) | 306 (25.9) |
| Social media is a useful tool for learning and accessing information. | 450 (38.0) | 525 (44.4) | 128 (10.8) | 54 (4.6) | 26 (2.2) |
| I am cautious about sharing personal information on social media. | 514 (43.4) | 397 (33.6) | 149 (12.6) | 70 (5.9) | 53 (4.5) |
| I have received support and encouragement from online communities or groups. | 215 (18.2) | 406 (34.3) | 271 (22.9) | 159 (13.4) | 132 (11.2) |
| I often feel overwhelmed by the constant stream of information on social media. | 242 (20.5) | 407 (34.4) | 292 (24.7) | 135 (11.4) | 107 (9.0) |
| Social media helps me discover new hobbies and interests. | 382 (32.3) | 539 (45.6) | 168 (14.2) | 47 (4.0) | 47 (4.0) |
| I have taken breaks from social media to improve my well-being. | 402 (34.0) | 389 (32.9) | 178 (15.0) | 119 (10.1) | 95 (8.0) |
| Social media can be a source of stress and anxiety for me. | 374 (31.6) | 386 (32.6) | 204 (17.2) | 129 (10.9) | 90 (7.6) |
| I am mindful of my screen time and try to balance it with other activities. | 353 (29.8) | 443 (37.4) | 210 (17.8) | 114 (9.6) | 63 (5.3) |
| I have used social media to seek advice or guidance on personal issues. | 352 (29.8) | 379 (32.0) | 192 (16.2) | 115 (9.7) | 145 (12.3) |

##### Box 1 What Moroccan Youth Identify as Key Drivers of Mental Health

The top 10 drivers that were given the highest ratings of importance by Moroccan youth, in descending order, were:

1. A good relationship with parents and family,
2. Access to essential needs such as food, water, a place to live, warmth, clothing, as well as feeling safe and secure,
3. A sense of purpose in life,
4. Feeling safe in daily life, whether at home, in the neighborhood, online, or at school or work,
5. Personal information is protected and is not shared without permission,
6. Having self-confidence and feeling capable to do things well,
7. Belief in the self and their ability to reach learning goals,
8. Physical health,
9. Not being exposed to violence,
10. Feeling independent and capable of making personal decisions.

In the first component, good health and optimum nutrition, 46.3% of participants rated sleeping at night to be extremely important to their wellbeing, 59.7% rated being physically healthy as extremely important, 53.1% identified having access to quality health services as extremely important, and 56.5% reported breathing clean air as extremely important to their wellbeing.

In the second component, connectedness, positive values, and contribution to society, participants rated 10 items under connectedness, positive values, and contribution to society. 75.7% identified having a relationship with parents and family as extremely important to their wellbeing and 42.5% rated having a positive relationship with peers and colleagues as very important.

In the third component, safety and a supportive environment, participants rated 7 items under safety and a supportive environment and 64.7% identified feeling safe in daily life as extremely important to their wellbeing. 70% reported having essential needs such as food, water, a place to live, warmth, clothing, feeling safe and secure as extremely important to their wellbeing. 63.7% identified having their personal information protected as extremely important to their wellbeing.

In the fourth component, learning, competence, education, skills, and employability, participants rated 8 items under learning, competence, education, skills, and employability. 54.4% rated getting to go to school and having chances to keep learning as extremely important to their wellbeing, 62.2% identified feeling self-confidence and feeling that they can do things well as extremely important to their wellbeing.

In the fifth component, agency and resilience, participants rated 6 items under agency and resilience. 59.4% rated feeling independent and capable of making their own decisions as extremely important to their wellbeing, 68.5% rated having a sense of purpose in their lives as extremely important to their wellbeing, and 55% rated having hope and optimism towards the future as extremely important.

Under the sixth component, digital wellbeing and literacy, participants rated 16 items using a scale from completely agree to strongly disagree. 37.7% of participants reported completely agreeing with social media helping them stay connected with friends and family and 48.2% agreed. 38% of participants rated social media as a useful tool for learning and accessing information as completely agree and 44.4% agreed. 43.4% selected completely agree and 33.6% agree in response to being cautious when sharing personal information on social media. 31.6% completely agreed and 32.6% agreed with social media being a source of stress and anxiety for them while 39% strongly disagreed with some people making fun of them on social media and affecting their wellbeing.

## 4. Discussion

The prevalence and social distribution of mental disorders is not as well documented in low to middle income countries, like Morocco, as it is in high income countries. Understanding the drivers of mental health and wellbeing in specific contexts allows for prevention efforts and the improvement of population health. This study sought to fill the gap of research on social and environmental drivers that impact the wellbeing and mental health of young people, from the perspective of young people’s lived experiences. Surveying Moroccan youth between the ages of 18 and 24 years old across the 12 regions of Morocco, using the five components of the Adolescent Wellbeing Framework, revealed the top ten drivers that Moroccan youth rated as highly important. The closer examination of the drivers given the highest rating of importance allows comparison with global mental health research on drivers and the identification of any specificities that warrant further research for the Moroccan youth population.

The results of this survey, under the first component of good health and optimum nutrition, and the second component of connectedness, positive values, and contribution to society, show that young Moroccans rate very highly the importance of positive relationships within their families and with parents as well as access to essential needs such as food, water, a home, clothing, and feeling safe. These results align with a systematic review of epidemiological studies that show a strong positive association between mental disorders and poverty in low to middle income countries (13), and studies showing that poorer mental health is found among populations who reported weak social support (14). A dominant hypothesis linking these drivers of low mental health is that low socioeconomic status may increase the level, frequency and duration of stressful experiences but these risk factors may be buffered with a good family relationship which is a source of social support (15). Therefore, for many participants in the study, who may be familiar with economic stressors, positive family and parental relationships rate higher as a priority slightly before basic needs. A study on the social determinants of mental health, conducted with youth in Indonesia, similarly highlights the importance of parental relationships with the young participants in the focus-group based study expressing that communication with parents and a sense of connection function as protective factors (16). These findings can provide evidence for programming to prioritize strengthening family relationships as a protective factor which can increase young people’s resilience in the face of adversity. Under the third and fourth components, safety and a supportive environment and learning, competence, education, skills, and employability, there were some interconnected findings. One of the determinants of poor mental health for young people is academic pressure and unsafe school environments (16–18) which pose an added stressor for young people and can contribute to accumulating stressors that impact one’s mental health. Participants in this study rated highly feeling a sense of purpose, self-efficacy, and capacity to learn as well as feeling safe and not being exposed to violence as important to their mental health. Education plays an important role in a range of later life outcomes, including employment, income, and community participation, but lack of support in learning or feeling unsafe in school due to bullying, can create a two-way relationship between struggling in school and feeling less confidence and self-efficacy (17,19–22). Literature from high income countries (23) as well as from other low-to-middle income countries (24) showcase a link between academic pressure and the way it impacts young people’s mental health, sense of purpose in life, confidence, and self-efficacy leading to poorer mental health outcomes.

Connecting the fifth component and sixth component, agency and resilience as well as digital wellbeing and literacy, young people rated privacy of personal information as highly important to them. Due to its implications for their ability to have agency and make their own life choices, this area of digital wellbeing requires further research to understand the impact of the changing digital landscape and the lack of a sense of privacy on the mental health and wellbeing of young people in low to middle income settings and collectivist cultures like Morocco.

The findings of this study reflect the lived experience of young people and their tacit awareness of the protective factors that help mitigate or buffer stressors and social determinants in their environment. They rate family relationships more highly than not being exposed to poverty and a sense of purpose and capacity to learn and do more highly than safety in schools. While all the drivers together paint a picture of what young people need to promote their mental health and positive development and the differences between the highly rated items may not be large enough for statistical significance, the results of this study can inform the priorities of intervention and programming. It highlights the need to strengthen the protective factors that matter to young people and make a difference in their mental health and resilience, especially when they may be exposed to stressors like poverty and unsafe school environments.

The study’s findings must be considered in light of several limitations. Firstly, the participants were identified through existing networks, potentially resulting in a sample that does not fully represent youth with differing abilities, needs, or socio-economic backgrounds. For instance, only 1.8% of respondents identified as NEET (Not in Education, Employment, or Training), indicating that the perspectives of this group may be underrepresented. Additionally, with only 14.3% of participants coming from rural contexts, the study may not fully capture the unique challenges and experiences of rural youth, who often face different socio-economic and infrastructural conditions compared to their urban counterparts. The survey used in this study was developed by the research team based on the Adolescent Wellbeing Framework which is a relatively new framework that is yet in the process of developing universal measures. As such, the lack of previous validation for this survey and its items introduces potential concerns regarding the internal and external validity of the findings.

Nevertheless, this study is the first of its kind in Morocco. It is a comprehensive study that collected data nationwide and centers the perspectives of young people. It provides important guidance for further research, policy, and practice. This study can help inform the training of counselors and mental health professionals (25) with local guidance on drivers of mental health to help address inequities which impact young people’s mental health. It can be used to guide youth programming, especially in providing interventions that support the development of positive relationships with parents and family members, that build a sense of purpose, self-efficacy, and capacity. It can also inform policy that seeks to address inequities through the reduction of poverty and creating safer environments for young people in schools, their communities, and at home. Having the perspectives of Moroccan youth from across 12 regions speaking to the drivers of mental health from their perspective can help enhance the inclusion of youth lived experiences in policy and practice and encourage further participatory research.

## Conclusions

This Nationwide Cross-Sectional study on the drivers of youth mental health and wellbeing addressed a key gap in literature by identifying important drivers of mental health and wellbeing among Moroccan youth between the ages of 18 and 24 years old across the 12 regions of Morocco. It is one of the first to use the Adolescent Wellbeing Framework (AWF) in data collection in an LMIC setting and expand on the framework’s five components with a sixth addressing digital wellbeing. The findings of this study included insightful differences in how young people in Morocco rate the drivers that matters most to them, which include the importance of family and parental relationships, having a sense of purpose, feeling safe at home, in their neighborhoods, and in school, and the protection of personal information online. These findings suggest that the definition of mental health and wellbeing and the drivers of wellbeing may be rated differently in a low to middle income setting in comparison to a high income setting which highlights the need for further research. The contribution of this study is significant in that it fills a large gap of data in the area of adolescent mental health in Morocco and in low to middle income countries and points to key priority areas for both research, intervention and policy development.

## Data Availability

All data produced in the present study are available upon reasonable request to the authors

## Abbreviations

LMICs: Low to middle-income countries
AWF: Adolescent Wellbeing Framework
NGO: Non-governmental organization
CSOs: Civil society organizations
NEET: Not in Education, Employment, or Training
MedSPAD: Mediterranean Survey on Alcohol and Other Drug Use in Schools.

## Declarations

### Ethics approval and consent to participate

The study protocol was reviewed and approved by the Ethics Committee of the Faculty of Medicine and Pharmacy of Tangier (AC470C/2023). Participation in the study was voluntary and uncompensated. Electronic informed consent was obtained from all participants prior to the beginning of data collection. All information collected from participants was kept confidential. This Study was conducted according to the guidelines of the Declaration of Helsinki.

### Consent for publication

Not applicable.

### Availability of data and materials

The data that support the findings of this study are available, from the corresponding author, upon reasonable request.

## Funding

This study was financially supported by Grand Challenges Canada.

### Authors’ contributions

MK conceptualized the study and supervised the work. OB, IEB, and MK were responsible for data curation, methodology development and investigation. IEB performed the formal analysis. JB wrote the original draft, and OB, IEB, and MK reviewed and edited the manuscript. All authors read and approved the final manuscript.

## Acknowledgments

The authors would like to thank the young people who participated in this study as well as the Being Initiative for their support and funding to implement this nationwide research project. The Being Initiative is a youth mental health fund hosted by Grand Challenges Canada, with partnership of Fondation Botnar, The UK’s National Institute for Health and Care Research (NIHR), Orygen, the Science for Africa Foundation, and United for Global Mental Health.

**Clinical trial number:** not applicable.

**Clinical trial number:** not applicable.

## References

1. World Mental Health Report: Transforming Mental Health for All. 1st ed. Geneva: World Health Organization; 2022. 1 p.

2. Being Initiative. Mapping Youth Mental Health Landscapes: Local Insights from 13 Countries [Internet]. Grand Challenges Canada; 2024. Available from: https://being-initiative.org/wp-content/uploads/2024/04/Mapping-Youth-Mental-Health-Landscapes_April-18-2024.pdf

3. Vereinte Nationen, editor. Youth social entrepreneurship and the 2030 agenda. New York, NY: United Nations; 2020. 1 p. (World youth report).

4. Conseil Économique, Social et Environnemental. La santé mentale et les causes de suicide au Maroc [Internet]. Rabat, Morocco: Conseil Économique, Social et Environnemental; 2023. Available from: https://www.cese.ma/media/2023/01/Rapport-sante%CC%81-mentale.pdf

5. Ministry of Health and Social Protection. The National Health Strategy for Adolescents and Youth 2022-2030. Rabat, Morocco: Ministry of Health and Social Protection; 2022.

6. Ministry of Education. Evaluation of the Use of Psychoactive Substances and Addictive Behavior among Students in Morocco (MedSPAD-IV MAROC 2021). Strasbourg, France: Council of Europe; 2021.

7. World Health Organization. Global School-Based Student Health Survey 2016 (MAR_2016_GSHS_v01) [Internet]. 2016. Available from: https://extranet.who.int/ncdsmicrodata/index.php/catalog/649

8. Baslam A, Kabdy H, Moubtakir S, Aboufatima R, Boussaa S, Chait A. Substance use among university students and affecting factors in Marrakech region, Morocco: a cross-sectional Study [Internet]. In Review; 2022 [cited 2025 Feb 12]. Available from: https://www.researchsquare.com/article/rs-1961768/v1

9. Zouini B, Sfendla A, Senhaji M, Råstam M, Kerekes N. Somatic health and its association with negative psychosocial factors in a sample of Moroccan adolescents. SAGE Open Medicine. 2019 Jan;7:2050312119852527.

10. Tom A, Mahfoud ZR. Factors associated with suicidality among school attending adolescents in morocco. Front Psychiatry. 2022 Aug 8;13:885258.

11. Molodynski A, Lewis T, Kadhum M, Farrell SM, Lemtiri Chelieh M, Falcão De Almeida T, et al. Cultural variations in wellbeing, burnout and substance use amongst medical students in twelve countries. International Review of Psychiatry. 2021 Feb 17;33(1– 2):37–42.

12. Ross DA, Hinton R, Melles-Brewer M, Engel D, Zeck W, Fagan L, et al. Adolescent Well-Being: A Definition and Conceptual Framework. Journal of Adolescent Health. 2020 Oct;67(4):472–6.

13. Lund C, Breen A, Flisher AJ, Kakuma R, Corrigall J, Joska JA, et al. Poverty and common mental disorders in low and middle income countries: A systematic review. Social Science & Medicine. 2010 Aug;71(3):517–28.

14. Lehtinen V, Sohlman B, Kovess-Masfety V. Level of positive mental health in the European Union: Results from the Eurobarometer 2002 survey. Clin Pract Epidemiol Ment Health. 2005;1(1):9.

15. Kelly Y, Sacker A, Del Bono E, Francesconi M, Marmot M. What role for the home learning environment and parenting in reducing the socioeconomic gradient in child development? Findings from the Millennium Cohort Study. Archives of Disease in Childhood. 2011 Sep 1;96(9):832–7.

16. Willenberg L, Wulan N, Medise BE, Devaera Y, Riyanti A, Ansariadi A, et al. Understanding mental health and its determinants from the perspective of adolescents: A qualitative study across diverse social settings in Indonesia. Asian Journal of Psychiatry. 2020 Aug;52:102148.

17. Abregú-Crespo R, Garriz-Luis A, Ayora M, Martín-Martínez N, Cavone V, Carrasco MÁ, et al. School bullying in children and adolescents with neurodevelopmental and psychiatric conditions: a systematic review and meta-analysis. The Lancet Child & Adolescent Health. 2024 Feb;8(2):122–34.

18. Khzami SE, Razouki A, Selmaoui S, Agorram B. Determinants of well-being of middle-school students in Moroccan urban and rural areas: A comparative study. J Edu Health Promot. 2020;9(1):271.

19. World Health Organization. Social determinants of mental health [Internet]. Geneva: World Health Organization; 2014 [cited 2025 Feb 13]. 52 p. Available from: https://iris.who.int/handle/10665/112828

20. Ye Z, Wu D, He X, Ma Q, Peng J, Mao G, et al. Meta-analysis of the relationship between bullying and depressive symptoms in children and adolescents. BMC Psychiatry. 2023 Mar 30;23(1):215.

21. Man X, Liu J, Xue Z. Effects of Bullying Forms on Adolescent Mental Health and Protective Factors: A Global Cross-Regional Research Based on 65 Countries. IJERPH. 2022 Feb 18;19(4):2374.

22. Pengpid S, Peltzer K. Prevalence and associated factors of psychological distress among a national sample of in-school adolescents in Morocco. BMC Psychiatry. 2020 Dec;20(1):475.

23. Kwak CW, Ickovics JR. Adolescent suicide in South Korea: Risk factors and proposed multi-dimensional solution. Asian Journal of Psychiatry. 2019 Jun;43:150–3.

24. Tran TV, Nguyen HTL, Tran XMT, Tashiro Y, Seino K, Van Vo T, et al. Academic stress among students in Vietnam: a three-year longitudinal study on the impact of family, lifestyle, and academic factors. J Rural Med. 2024;19(4):279–90.

25. Woods-Jaeger B, Cho B, Briggs EC. Training psychologists to address social determinants of mental health. Training and Education in Professional Psychology. 2024 Feb;18(1):31–41.

